# Positive correlation between long term emission of several air pollutants and COVID-19 deaths in Sweden

**DOI:** 10.1101/2020.12.05.20244418

**Authors:** Lars Helander

## Abstract

Several recent studies have found troubling links between air pollution and both incidence and mortality of COVID-19, the pandemic disease caused by the virus SARS-CoV-2. Here, we investigate whether such a link can be found also in Sweden, a country with low population density and a relatively good air quality in general, with low background levels of important pollutants such as PM2.5 and NO_2_. The investigation is carried out by relating normalized emission levels of several air pollutants to normalized COVID-19 deaths at the municipality level, after applying a sieve function using an empirically determined threshold value to filter out noise. We find a fairly strong correlation for PM2.5, PM10 and SO_2_, and a moderate one for NO_x_. We find no correlation neither for CO, nor (as expected) for CO_2_. Our results are statistically significant and the calculations are simple and easily verifiable. Since the study considers only emission levels of air pollutants and not measurements of air quality, climatic and meteorological factors (such as average wind speeds) can trivially be ruled out as confounders. Finally, we also show that although there are small positive correlations between population density and COVID-19 deaths in the studied municipalities (which are for the most part rural and non densely populated) they are either weak or not statistically significant.

## 1 Introduction

Air pollution is a global health concern and numerous studies have found links between air pollution and both respiratory and chronic diseases [1, 2, 3, 4]. Since the start of the COVID-19 pandemic, several studies have found links between air pollution and COVID-19 incidence and mortality [5, 6, 7, 8, 9]. The present study aims at determining whether there is a link between air pollution and COVID-19 deaths at the municipality level in Sweden. We propose that Sweden is a suitable candidate for this kind of ecological study, due to its long tradition of high quality statistics coupled with the fact that the COVID-19 pandemic has not been met here with severe governmental mitigation measures such as lockdown or school closures or other coercive measures such as obligations to wear face masks. Also, regional responses to the pandemic have not varied much throughout the country, thus encouraging the disease to run its natural course and reach all corners of the country.

The method of the present study, which will be detailed in Section 2, consists in a first step of relating normalized emission levels to normalized COVID-19 deaths at the municipality level. Normalization refers here to relating the figures to the number of inhabitants of each municipality. A positive correlation thus means that municipalities with relatively high emissions and few inhabitants will account for relatively more COVID-19 deaths than those with lower emissions or more inhabitants. In Sweden, municipalities with high emissions and few inhabitants are traditionally called “bruksorter” (mill towns), and are centered around one or several industries (paper mill, steel mill, factory, mine, etc.) playing a major role in the lives of the inhabitants. The industries are required by law to report yearly emission levels of hazardous substances to a public register.

In a second step, we introduce a threshold value *t*_*q*_ for each pollutant *q*. Municipalities with normalized emissions of *q* below the threshold value *t*_*q*_ are excluded from the calculation of the correlation coefficient. The rationale for using a threshold is to remove noise, as COVID-19 death figures in municipalities with low emissions are likely to be driven by other factors than those emissions. Municipalities with high emissions and many inhabitants (such as big cities) may thus fall under the threshold value *t*_*q*_ even though the emissions very well may have an impact on COVID-19 mortality. However, excluding those cases should not lower the relevance of the study’s results.

## 2 Method

Emission [10], census [11] and population density [11] data at the municipality level were retrieved directly from Statistics Sweden (SCB), the Swedish government statistics agency with a history dating back to before 1749.

COVID-19 deaths registered up to November 30, 2020 [12] at the municipality level were retrieved directly from the Swedish National Board of Health and Welfare (Socialstyrelsen), another government agency. No data for municipalities with 1-3 COVID-19 deaths is given in [12] due to privacy concerns (in this case an ‘X’ is given in the table instead). For those municipalities, we assume for simplicity that there are 2 COVID-19 deaths.

Now, let *q* denote the pollutant under study, with *q* ∈ {`PM2`.`5`, `PM10`, SO_2_, NO_x_, `CO`, CO_2_}. Let *M* denote the set of municipalities in Sweden (|*M* | = 290). For each *m* ∈ *M*, let 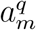 denote the average emission of *q* in *m* for the years 2008 to 2018 (inclusive), let *p*_*m*_ denote the number of inhabitants of *m* for the year 2019 and let *c*_*m*_ denote the number of COVID-19 deaths in *m* up to November 30, 2020.

The normalized emissions 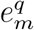 of *q* in *m* is calculated according to:

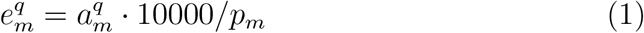

The normalized COVID-19 deaths *d*_*m*_ in *m* is calculated according to:

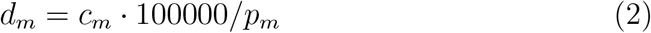

The following threshold values *t*_*q*_ for each type of pollutant has been identified empirically using the available data. (No correlation has been discernible for `CO` or CO_2_ for any threshold):

We now introduce the sets *S*_*q*_ per pollutant, representing our samples, with the following definition (using the sieve function 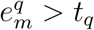 to filter out noise):

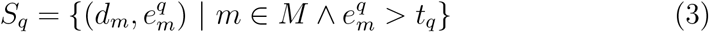

For each pollutant *q* we compute Pearson’s correlation coefficient *r* and *p*-value of our sample *S*_*q*_ and a 95% confidence interval in the standard way using [13].

To determine any possible relationship between population density and COVID-19 deaths, we let *ρ*_*m*_ denote the number of inhabitants per square kilometer in the municipality *m* and we define the set *P* and sets *P*_*q*_ for each pollutant *q* as follows:

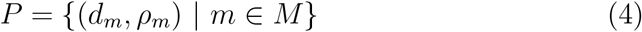

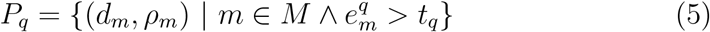

As a comparison, we also define the set 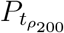 below, which corresponds to the set of municipalities with a population density above 200 inhabitants per square kilometer. The threshold of 200 has been empirically determined, in the same way as the thresholds for the pollutants.

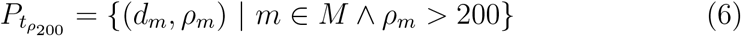

We then compute Pearson’s correlation coefficient and 95% confidence interval in the same way as before for all these sets (using [13]).

## 3 Results

Below in Table 2 follows the main results of our study. For scatter plots, please refer to Appendix A.

**Table 1:** Table of empirically determined threshold values for each pollutant.

| *Pollutant* ($q$) | *Threshold* ($t_q$) | *Unit* |
| --- | --- | --- |
| PM2.5 | 60 | tonnes per 10000 inhabitants |
| PM10 | 100 | tonnes per 10000 inhabitants |
| SO<sub>2</sub> | 70 | tonnes per 10000 inhabitants |
| NO<sub>x</sub> | 220 | tonnes per 10000 inhabitants |
| CO | 0 | tonnes per 10000 inhabitants |
| CO<sub>2</sub> | 0 | 1000 tonnes per 10000 inhabitants |

**Table 2:**
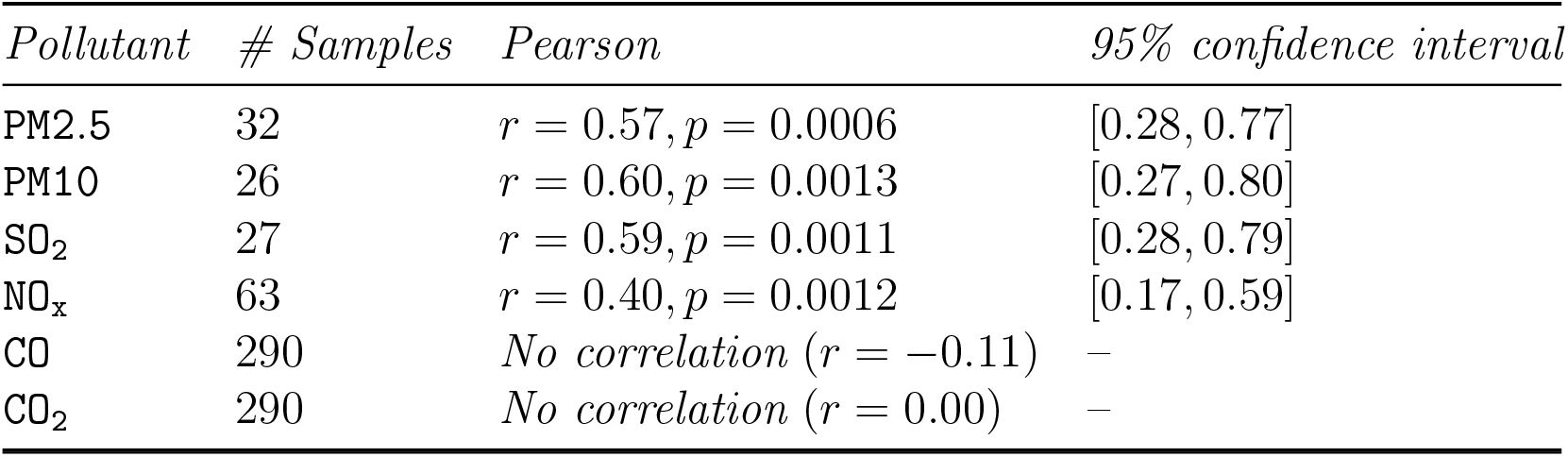
Pearson’s correlation coefficient and 95% confidence interval for the set *S*_*q*_ for each pollutant *q*, representing the relationship between emission levels of *q* and COVID-19 deaths, as defined in Section 2.

In Table 3 below, we present the correlations between population density and COVID-19 deaths in the different sets of municipalities.

**Table 3:** Pearson’s correlation coefficient and 95% confidence interval for the set *P* and sets *P*_*q*_ for each pollutant *q*, representing the relationship between population density and COVID-19 deaths, as defined in Section 2. 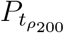 corresponds to the set of municipalities with a population density above 200 inhabitants per square kilometer.

| *Population density* | *# Samples* | *Pearson* | *95% confidence interval* |
| --- | --- | --- | --- |
| $P_{\text{PM2.5}}$ | 32 | $r = 0.24, p = 0.18$ | [−0.12, 0.55] |
| $P_{\text{PM10}}$ | 26 | $r = 0.22, p = 0.28$ | [−0.18, 0.56] |
| $P_{\text{SO}_2}$ | 27 | $r = 0.10, p = 0.63$ | [−0.29, 0.46] |
| $P_{\text{NO}_x}$ | 63 | $r = 0.28, p = 0.02$ | [0.04, 0.50] |
| $P$ | 290 | $r = 0.22, p = 0.0001$ | [0.11, 0.33] |
| $P_{t_{\rho_{200}}}$ | 28 | $r = 0.46, p = 0.01$ | [0.12, 0.70] |

## 4 Conclusions

After applying a sieve function to filter our noise, we have proved statistically significant correlations between the air pollutants PM2.5, PM10, SO_2_ and NOx and COVID-19 deaths at the municipality level in Sweden. The correlations for the three first substances are fairly strong, and for the last the correlation is more moderate. Our approach is new in the sense that we consider only emission levels of pollutants and not measurements of air quality, which means that we can easily exclude climate and weather (such as wind speeds) as confounding factors. We have also shown that the correlations between population density and COVID-19 deaths in the studied municipalities are either weak or not statistically significant, which means that the observed correlations with air pollution are not due to population density. When considering the country as a whole, there is a weak, but positive, correlation between population density and COVID-19 deaths. If we restrict ourselves to the municipalities with a population density above 200 inhabitants per square kilometer, the correlation becomes more pronounced.

### Limitations

A limitation to this ecological study is that we cannot quantify to what extent toxic substances emitted in a municipality are actually inhaled by its inhabitants and thus to what extent they play a relevant role in the observed correlations. As a future improvement, we would like to incorporate meteorological factors at the municipality level, in particular average wind velocities, into our calculations as a means to modelize the speed of dispersal of the polluting substances. Also, we would like to remind the reader that we have only proved a correlation between data in statistical tables, not any causal relationship between air pollution and COVID-19 deaths. Nevertheless, we hope that this novel approach can be of value to the scientific community and to policymakers, as another small piece in the puzzle of the relationship between air pollution and disease burden in our societies.

## Data Availability

All data is freely available on the Internet.

https://www.statistikdatabasen.scb.se/pxweb/en/ssd/START__BE__BE0101__BE0101C/BefArealTathetKon/

https://www.socialstyrelsen.se/globalassets/1-globalt/covid-19-statistik/statistik-over-antal-avlidna-i-covid-19/statistik-covid19-avlidna.xlsx

## A Scatter Plots

**Figure 1:**
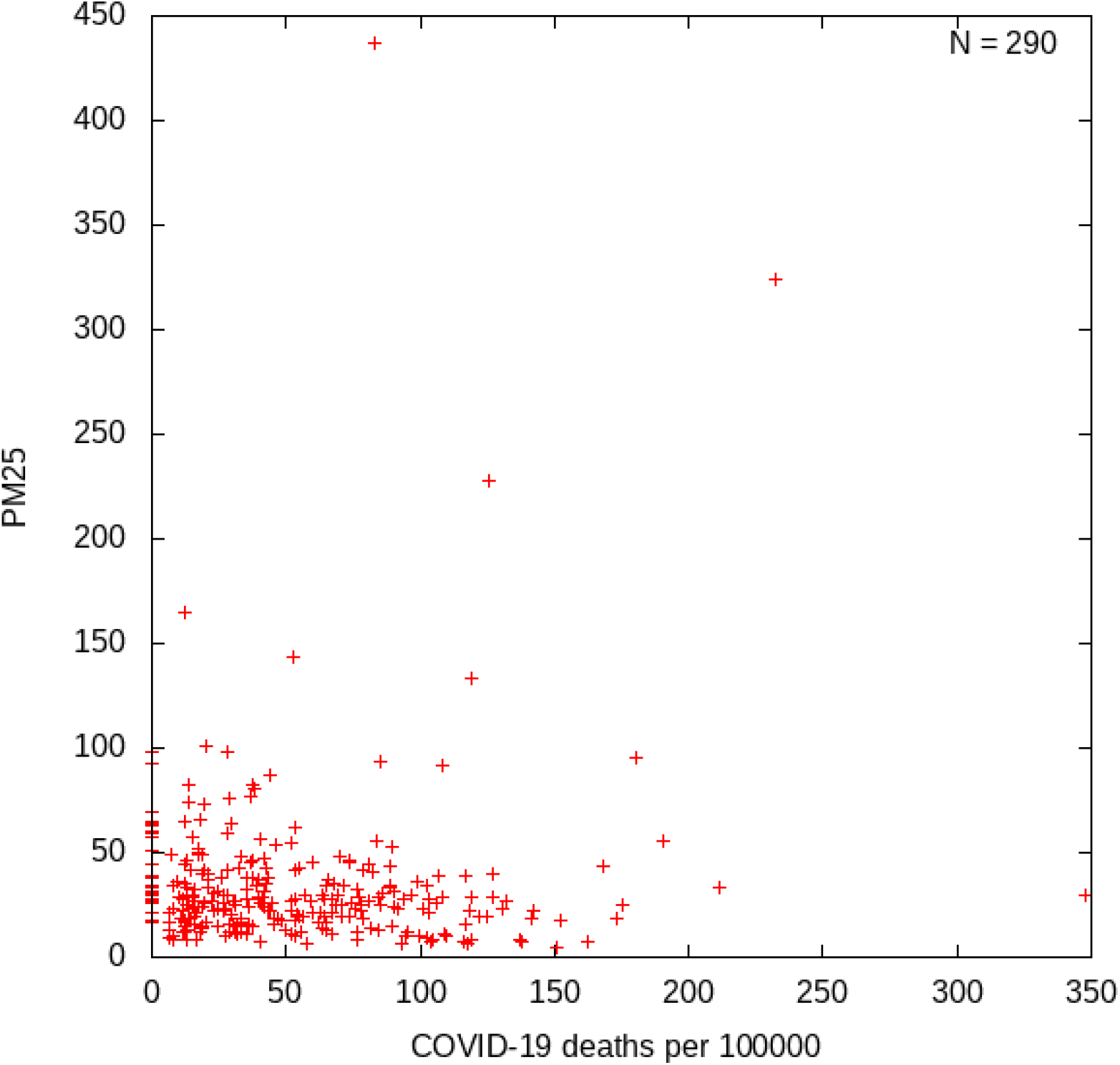
Emissions of PM2.5 (tonnes per 10000 inhabitants) related to COVID-19 deaths. One point per municipality.

**Figure 2:**
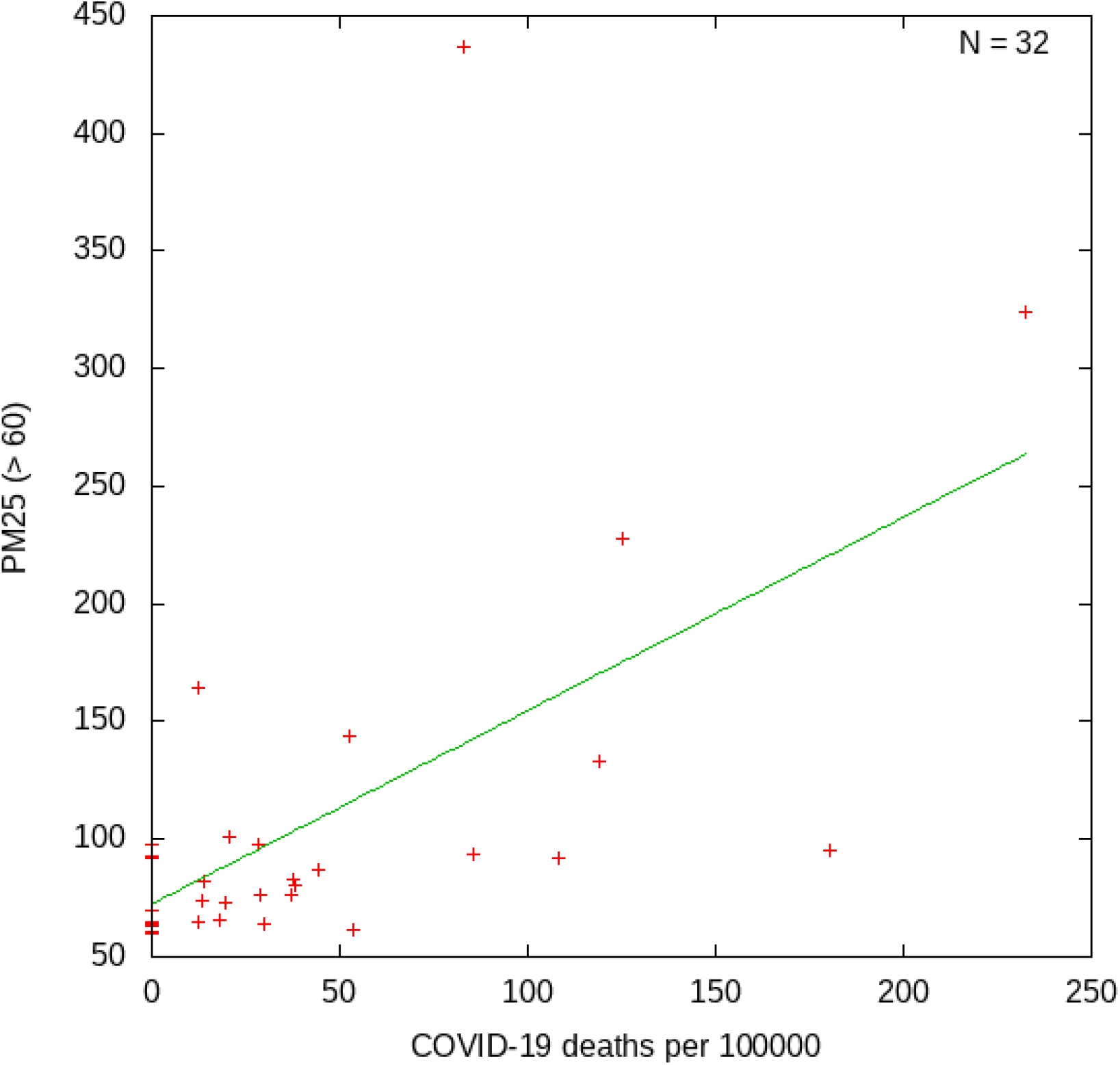
Emissions of PM2.5 (tonnes per 10000 inhabitants) related to COVID-19 deaths. One point per municipality with emissions *> t*_PM2.5_. The included municipalities are Arjeplog, Askersund, Bengtsfors, Dorotea, Gotland, Grums, Gällivare, Härjedalen, Kalix, Karlshamn, Kiruna, Kramfors, Kristinehamn, Lindesberg, Lysekil, Mönsterås, Norsjö, Oxelosund, Piteå, Ragunda, Robertsfors, Rättvik, Sorsele, Storuman, Tjörn, Vilhelmina, Vindeln, Ydre, Åsele, Ä lvkarleby, Örnskoldsvik, Överkalix.

**Figure 3:**
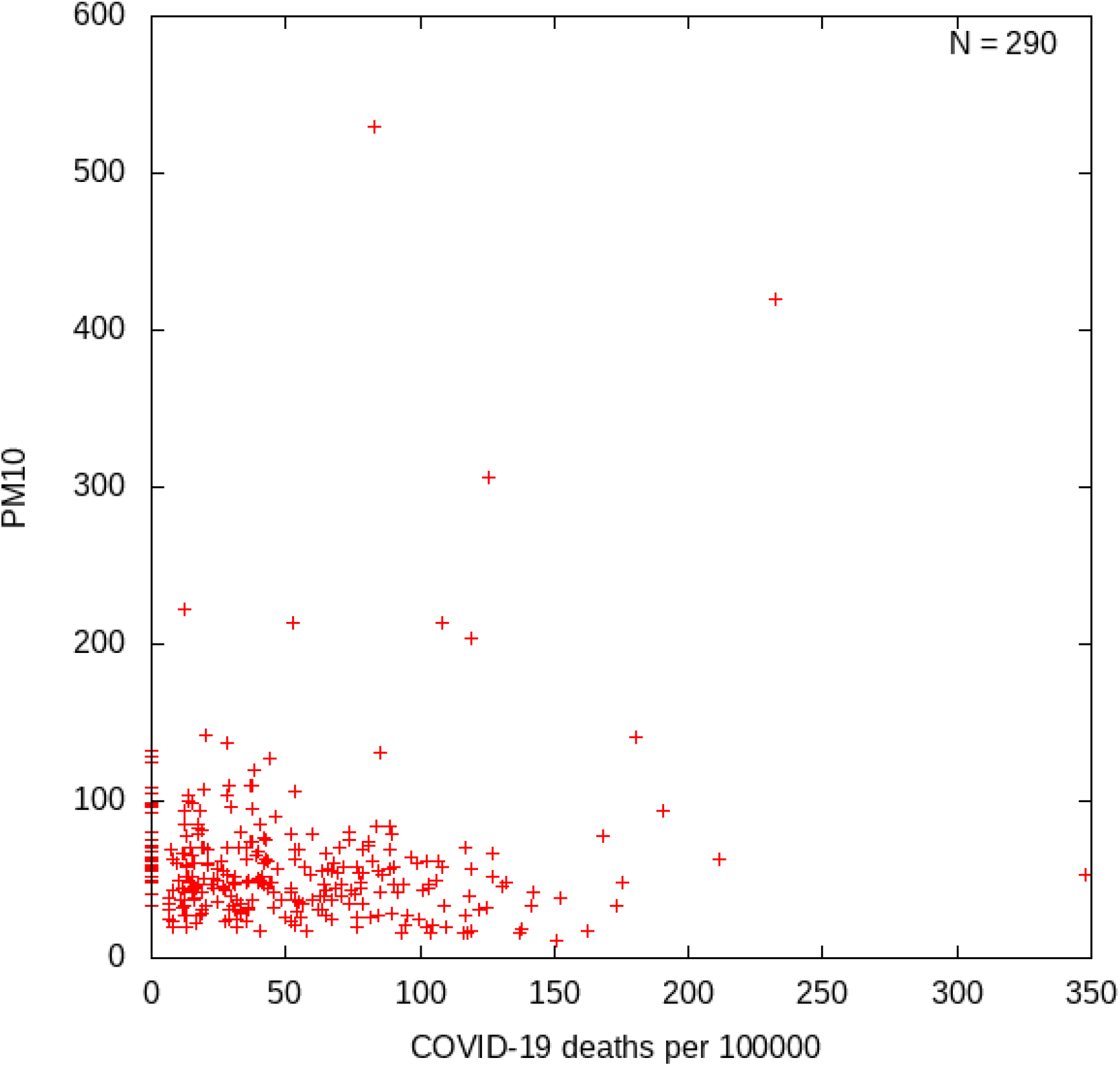
Emissions of PM10 (tonnes per 10000 inhabitants) related to COVID-19 deaths. One point per municipality.

**Figure 4:**
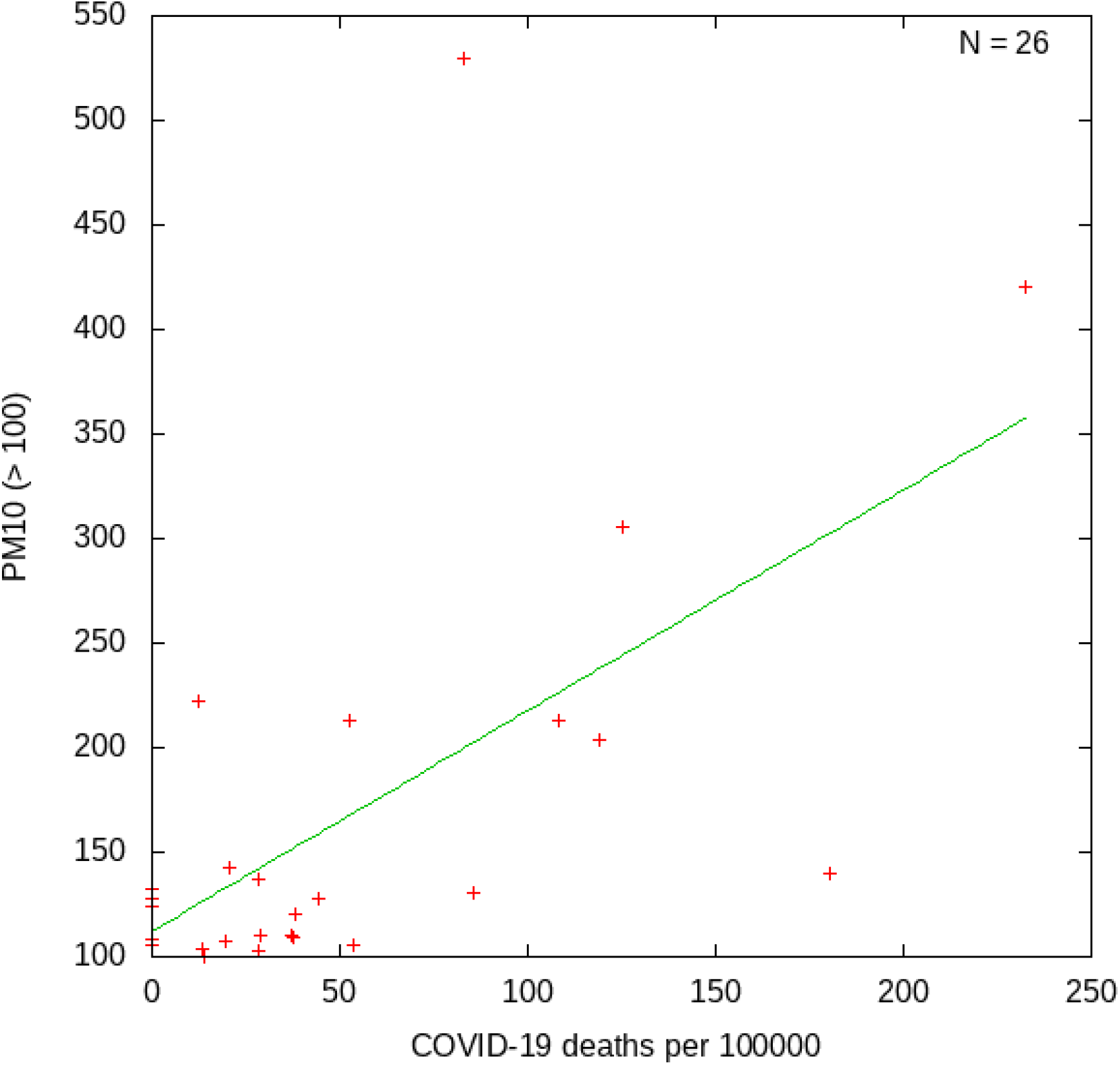
Emissions of PM10 (tonnes per 10000 inhabitants) related to COVID-19 deaths. One point per municipality with emissions *> t*_PM10_. The included municipalities are Askersund, Bengtsfors, Berg, Dorotea, Gotland, Grums, Gällivare, Kalix, Kiruna, Kramfors, Kristinehamn, Lindesberg, Lysekil, Mönsterås, Norsjö, Oxelösund, Piteå, Ragunda, Sorsele, Storuman, Tjörn, Vilhelmina, Vindeln, Ydre, Älvkarleby, Örnskoldsvik.

**Figure 5:**
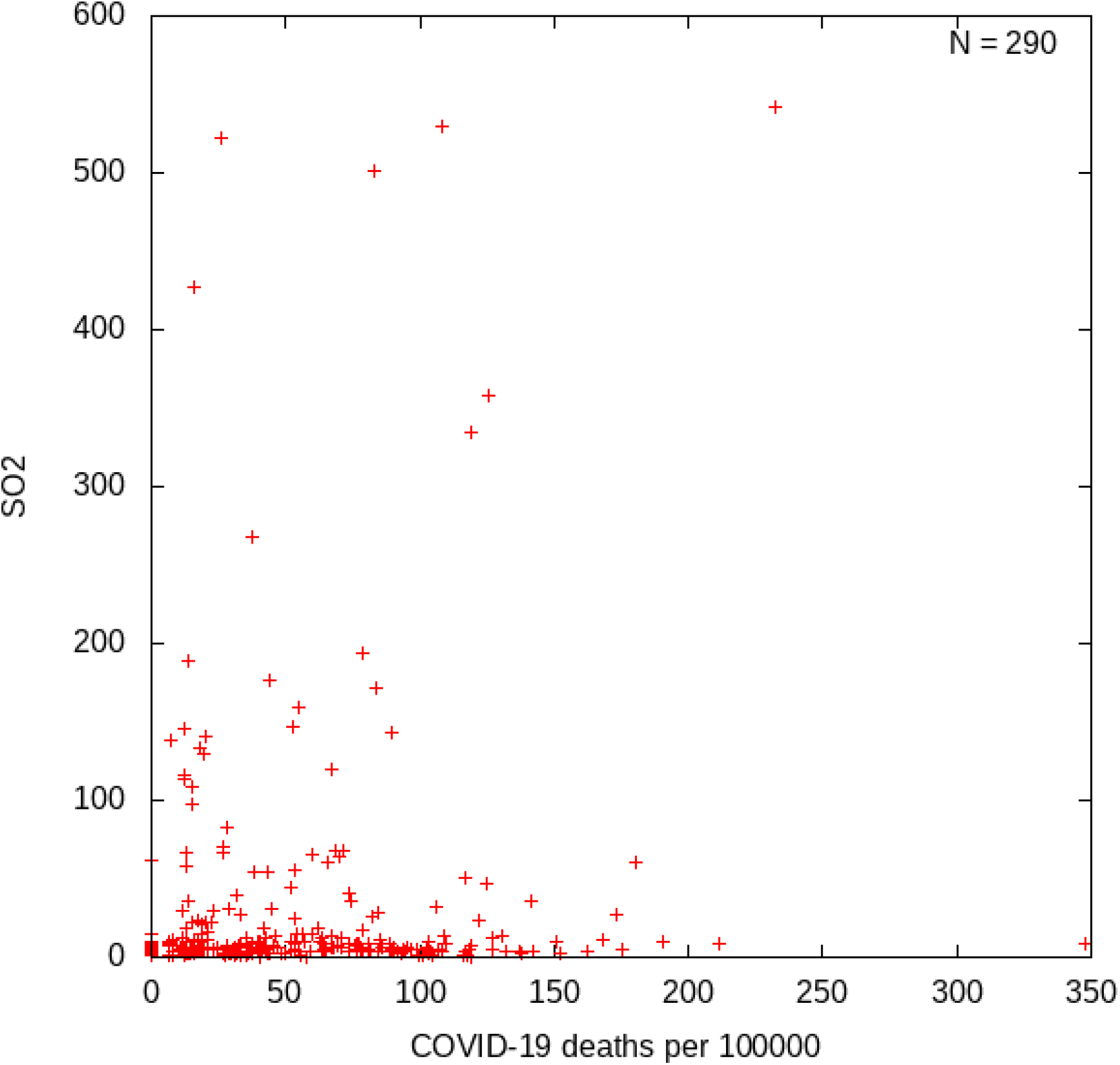
Emissions of SO_2_ (tonnes per 10000 inhabitants) related to COVID-19 deaths. One point per municipality.

**Figure 6:**
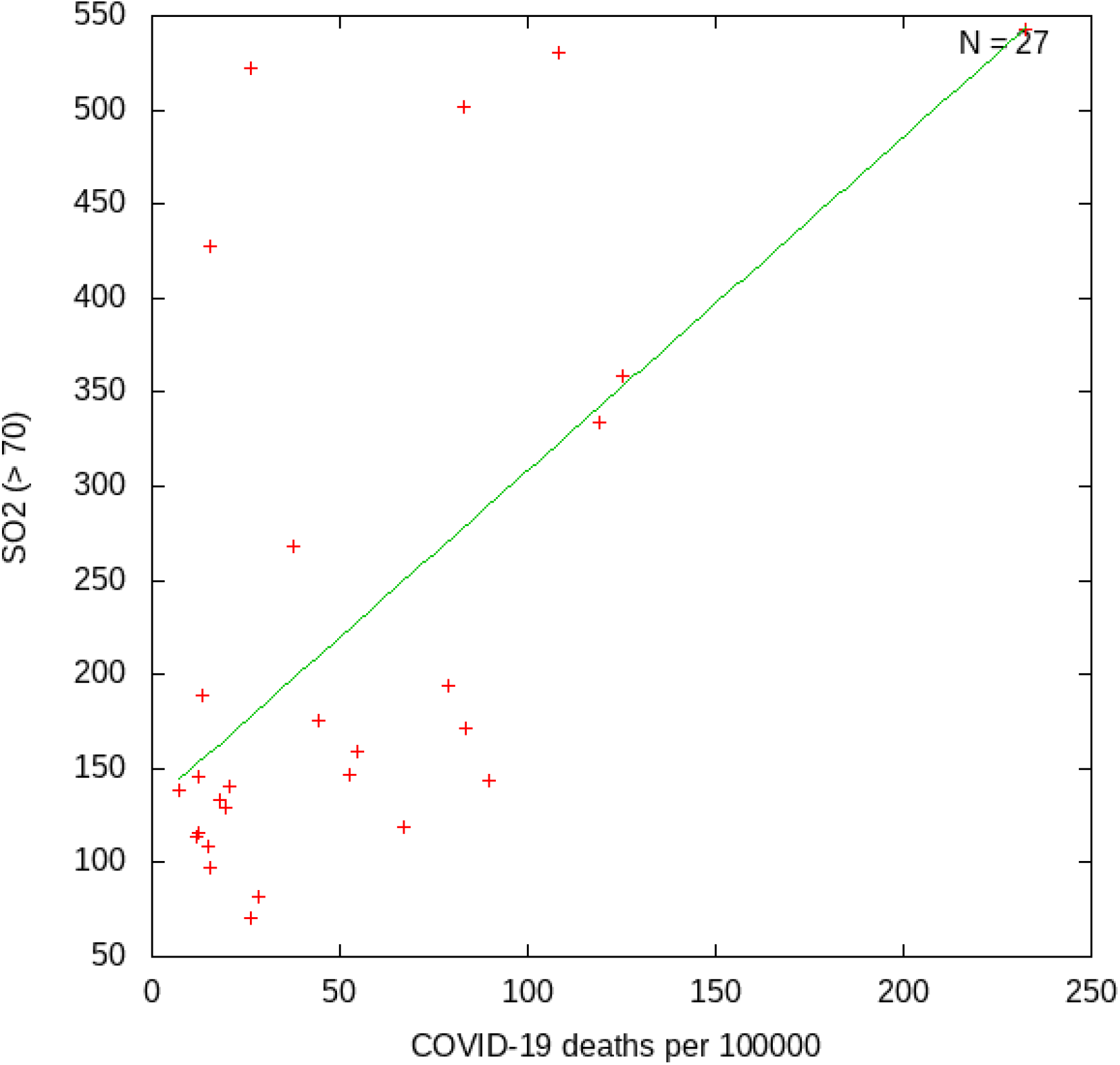
Emissions of SO_2_ (tonnes per 10000 inhabitants) related to COVID-19 deaths. One point per municipality with emissions 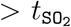. The included municipalities are Askersund, Bengtsfors, Bromölla, Gotland, Grums, Gällivare, Göteborg, Götene, Hammarö, Kalix, Karlshamn, Kiruna, Lidköping, Luleå, Mönsterås, Oxelösund, Piteå, Rattvik, Skellefteå, Stenungsund, Söderhamn, Timrå, Tjörn, Tranemo, Trelleborg, Älvkarleby, Örnskoldsvik.

**Figure 7:**
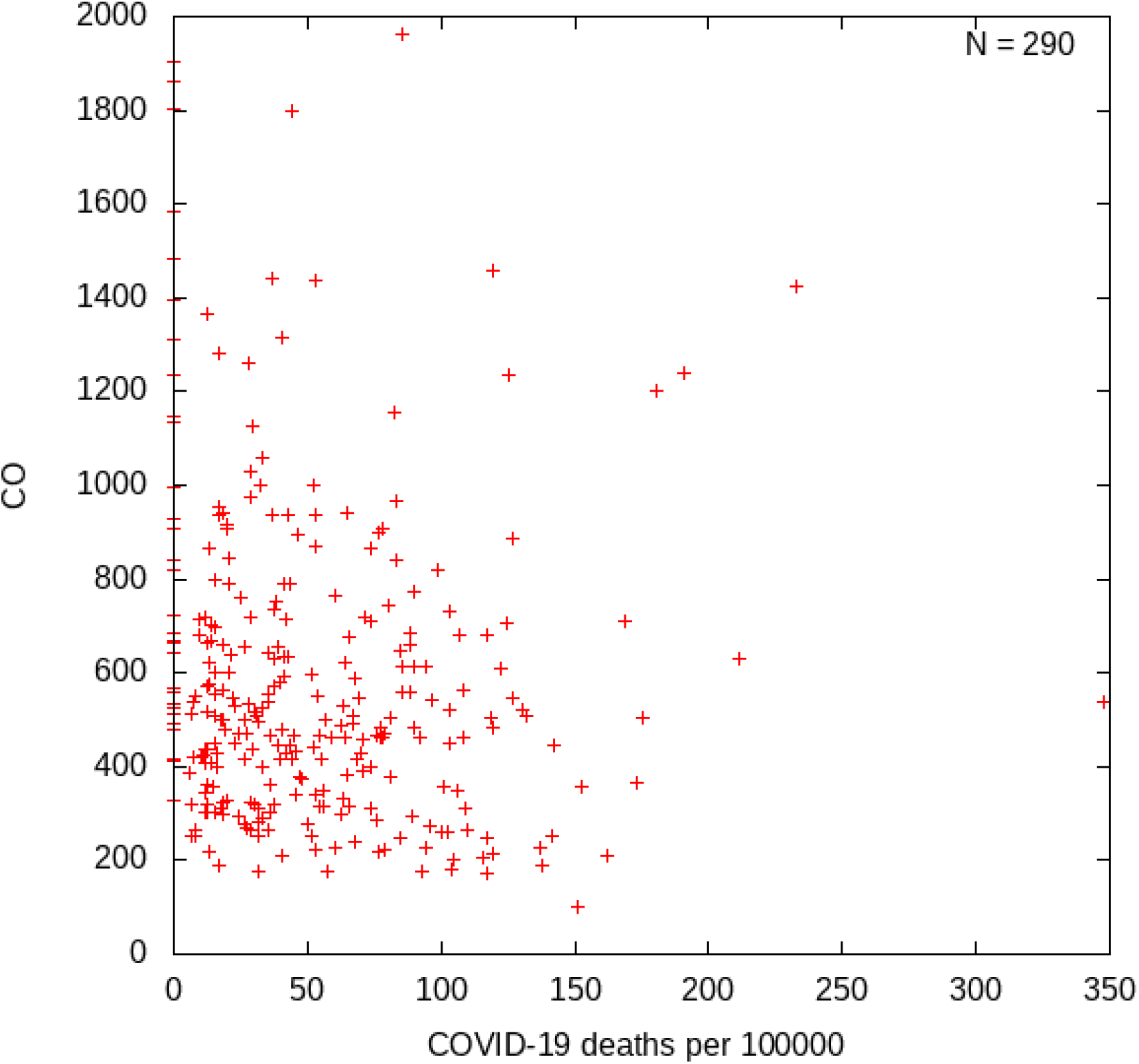
Emissions of CO (tonnes per 10000 inhabitants) related to COVID-19 deaths. One point per municipality. *No correlation found*.

**Figure 8:**
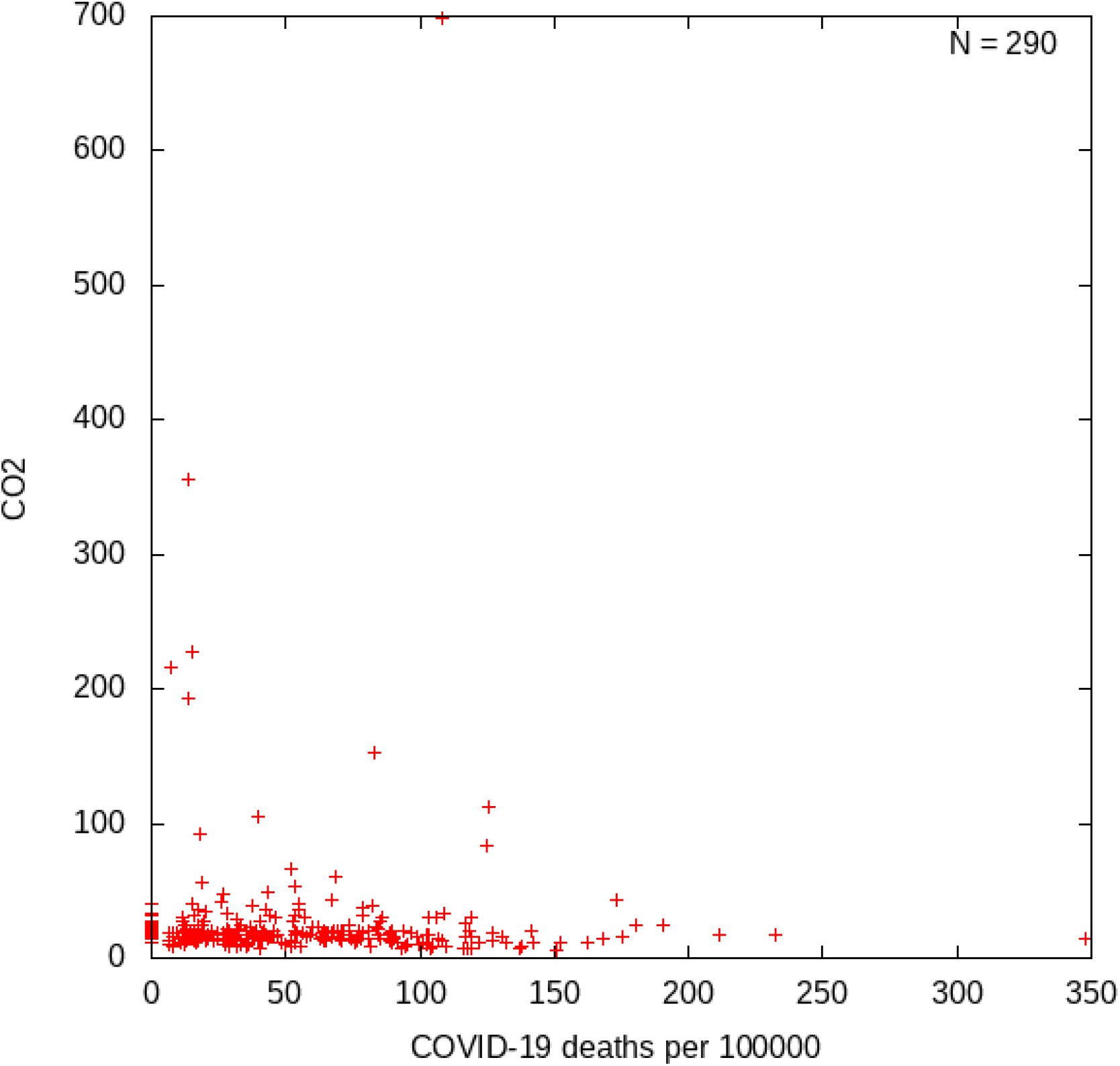
Emissions of CO2 (1000 tonnes per 10000 inhabitants) related to COVID-19 deaths. One point per municipality. *No correlation found*.

**Figure 9:**
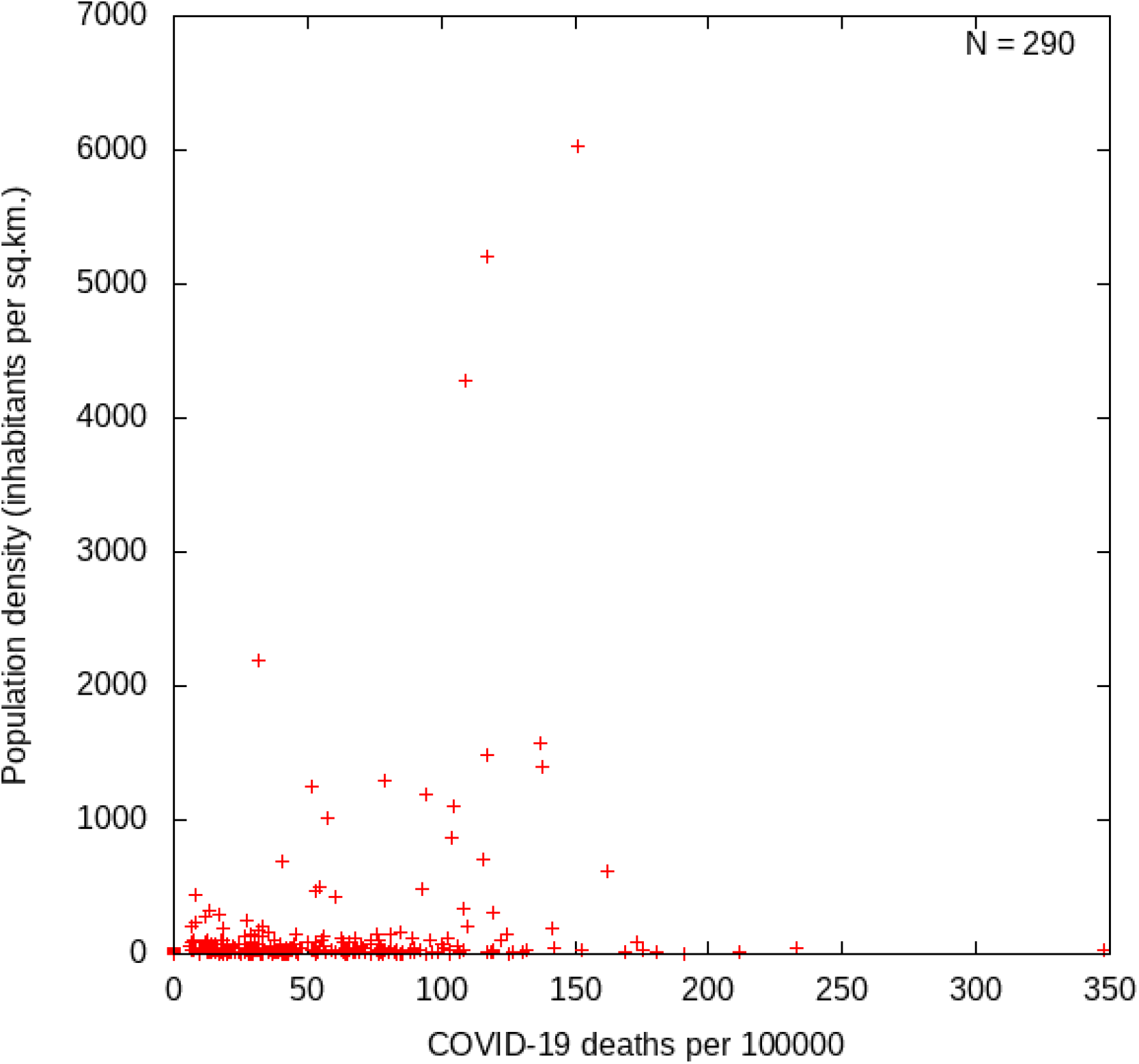
Population density (inhabitants per square kilometer) related to COVID-19 deaths. One point per municipality.

## Notes

### Competing Interest Statement

The authors have declared no competing interest.

### Funding Statement

There is no funding of this work.

### Author Declarations

This is independent research, no approvals seem necessary, but perhaps I am mistaken.

